# Causal Inference for Estimation of Vaccine Effects from Time-to-Event Data

**DOI:** 10.1101/2023.09.24.23296040

**Authors:** Zhouxuan Li, Kai Zhang, Yashar Talebi, Hulin Wu, Wenyaw Chan, Eric Boerwinkle, Momiao Xiong

## Abstract

Vaccine is the most efficient method for controlling of infectious disease. Vaccine effectiveness estimation is extremely important in monitoring vaccine efficacy and controlling disease spreading. To study about the COVID-19 vaccine effectiveness from EHR data, we apply the counterfactual reasoning method with deep neural network for vaccine effectiveness estimation from the time-to-event data which are extracted from Optum EHR dataset. The estimated vaccine effectiveness by the counterfactual reasoning is compared with the Cox regression model and Random survival forest model. The preliminary results show that the proposed model is more unbiased than the Cox regression and Random survival forest models.

## Introduction

Until May 2023, the COVID-19 pandemic had led to 160 million cases and 1.14 million deaths in the United States of America [1]. The spread of the severe acute respiratory syndrome coronavirus 2 (SARS-CoV-2) has been slowed down after the wide injection of vaccines. The effect of vaccination for preventing disease is a treatment effect estimation problem. Many studies are conducted to study the efficacy of COVID-19 vaccinations [2-7], and most of them are strictly designed clinical trials. In this project, the EHR data from Optum COVID-19 dataset will be used to estimate COVID-19 vaccination efficacy for preventing infection. The causal inference by counterfactual reasoning method would be applied as the analysis framework.

Vaccine effectiveness (VE) estimation can be considered as a treatment effect estimation problem in which the vaccine is a treatment against a particular virus or pathogen infection. Vaccine effectiveness (VE) estimation has been a key study area for infectious diseases. Accurate estimation of the extent of the waning of vaccine-induced protection over time is an important public health need [8]. Vaccine efficacy (VE) is generally estimated by:*VE* = 1 − *RR*, where *RR* is the measurement of relative risk in the vaccinated compared with the unvaccinated group. The time-varying VE will be estimated by *VE* (*t*) = 1 – *RR* (*t*), where *RR* (*t*) is the rate ratio at a specific time point *t* [9]. When estimating vaccine effectiveness in observational studies, scientists often encounter the problems of selection bias and covariate shift. Selection bias is generated when patients get their treatment by nonrandomized procedures. It will make observational data distributed differently from the population under research. When the observational covariate distribution differs from the whole population, it becomes a covariate shift problem. In our research, we adapt the model with time-to-event data in a counterfactual causal inference framework (survITE) [10] for vaccine effectiveness estimation. The VE estimation is conducted within EHR dataset, and compared with the results from a Cox regression model [11] and Random survival forest (RSF) model [12].

## Methods and material

### Problem formulation

The effectiveness of vaccination can be formulated as a problem of estimation of the effects of the treatment on survival time. We consider a vaccine as a treatment, denoted as a = 1 or 0 where a = 1 denotes vac cine inoculated, and a = 0 denotes vaccine uninoculated. Let *A* be the treatments vector in observational data. Consider the elapsed time during which one has not been infected by an infectious disease such as COVID-19 after their inoculation as survival time, denoted byτ, and *T* be the time vector in observational data. Let *X* be a covariate vector associated with a patient’s characteristics and features, and *C* be a censoring time which indicates loss of follow-up for a patient. Therefore, the observed survival time vector 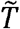 (the time elapsed until either breakthrough infection or censoring occurs) is defined as

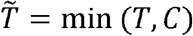

We define indicator variable Δ = *I*(*T* ≤ *C*), which shows whether the breakthrough infection is obtained or not. Consider *n* patients. The observed dataset is

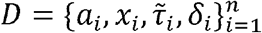

where 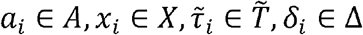.

### Conditional Hazard

To define the conditional hazard function *λ*(τ | *a, x*)we need to track breakthrough infection and censoring events. Define the indicators at the time *t* for the breakthrough infection and the censoring event, respectively, as 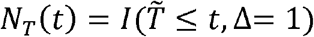 and 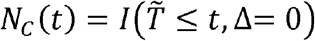 for *t* ∈ *T*. Define event indicator at the time *t* as *Y*(*t*) = *I* (*N*_*T*_ (*t*) = 1 ∩ *N*_T_ (*t* – 1) = 0), i.e., the event occurs exactly at time *t* The conditional hazard at the time τ is defined as the conditional probability, which means an event occurs at time τ and it does not occur before time τ [10]:

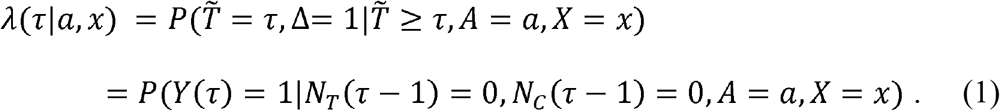

We define the risk set at the time τ as

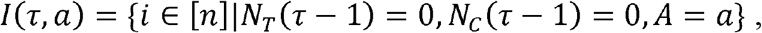

i.e., the set of samples at risk at time τ.

The survival function at time τ is defined as

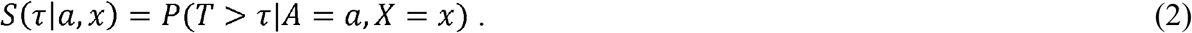

The survival time and censoring time probability functions are defined, respectively, as

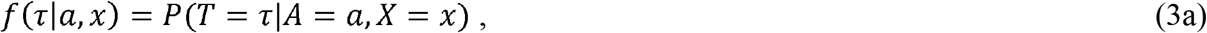

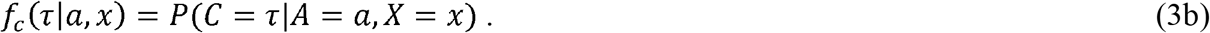

Then, the conditional hazard λ (τ|*a, x*) can be expressed as

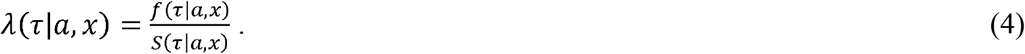

The relationship between the hazard function and survival function is given by

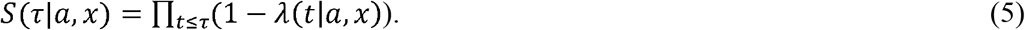

### Conditional Hazard under Intervention and Estimation of the Vaccine Effectiveness

The above hazard and survival functions are defined in terms of association. The treatment effect estimation involves causal analysis. The hazard, survival functions, and treatment effect estimation should be defined in terms of interventions. Let *do* (*A* = *a*) denote do-operator, implying that every individual is randomly assigned to a treatment *a*. Then, the hazard and survival functions under intervention are defined as [10] :

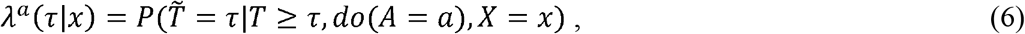

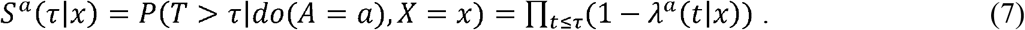

The vaccine efficiency at day, is defined as the proportional reduction in the hazard rate of infection after τ day vaccination for an individual [2]:

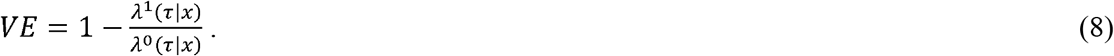

Equation (8) implies that vaccine efficiency estimation needs to estimate counterfactual hazards. These are essential quantities in cause-based treatment effect estimation from survival data. To identify these individual quantities, we make some assumptions for the data [10]:

1. Data are generated by general directed acyclic graphs (DAGs) (supplementary Figure 1) where a set of covariates are divided into possibly overlapping subsets *X =* {*X*_1_, *X*_2,_ *X*_3_}, represents a treatment. The DAG also includes infection and censoring events at time *t*_1_, *t*_2_,…,*T*.
2. Assume that there are no hidden confounders in static treatment effect estimation, i.e., *T*_*a*_ *A* | *X*.
3. Assume that censoring is random, i.e.,*T*_*a*_ *C* | *X, A*.
4. Consistency assumption. Assume that the observed outcomes are potential outcomes under the observed intervention, i.e., λ^*a*^ (τ | *x*) = λ (τ | *x*).
5. Overlap/positivity. Assume that with non-zero probability(*P >* 0), the interventions of interests are observed, i.e.,0 < ε_1_, ε_2_ ε_3_ < 1, for we assume:
  1. *ε*_1_ < *P(A* = *a|X = x*) < 1 − *ε*_1_
  2. *P(N*_*c*_(*t*) = 0|*A* = a, *X = x) = P(C>t*|*A = a, X = x> ε*_*2*_ *for all t<T*
  3. *P(N*_*T*_ *(t - 1) = 0*|*A = a, X = x) = P(T > t − 1*|*A = a, X = x) > ε*_3_

To estimate hazard, we consider event likelihoods and censoring separately since censoring is random and ignorable. The likelihood contribution of observation *i* for hazard estimation is given by [10, 13].

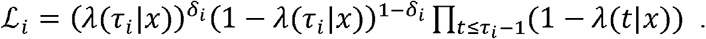

The summation of negative log-likelihood is

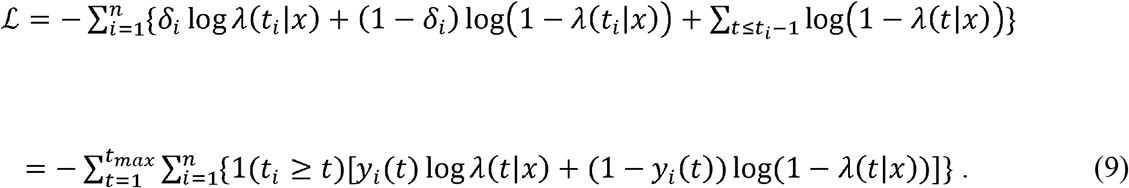

Let *P*(*T* > *t* | *X* = *x*) = *E*[1(*T* > *t* | *X = x*)] = *S* (*t* | *x*)There are two possibilities:*T > t* and *T* ≤ *t* When *T* ≤ *t*, it indicates that is not censored. We need to specify *δ*_*i*_ = 1 Therefore, the survival-based negative log-likelihood is given by:

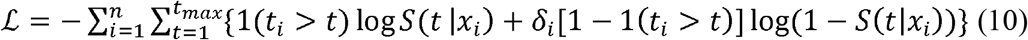

Finally, we consider the probability mass function (PMF) - based estimation. By definition of hazard rate, we obtain that

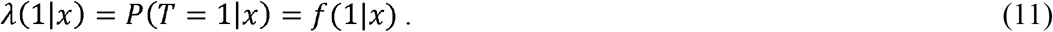

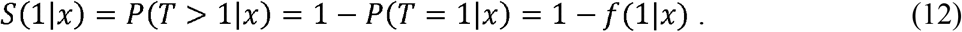

Assume one-hot encoded label [1 (*t*_*i*_ = *t*)]_*t*∈ 𝒯_. Then, we have

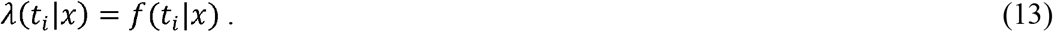

Using equations (9) and (13), we obtain the PMF-based negative log-likelihood:

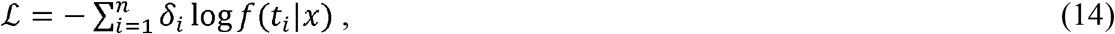

where each uncensored observation contributes to this likelihood.

Define the stochastic time-to-event (infection) generative function *h*_*ar*_ (.) Assume that potential outcomes *t*_*a*_ are sampled from the distribution 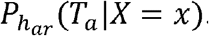. Let 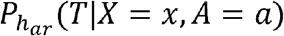 be an event time distribution. Then, the negative log-likelihood loss function *l*(*Y*(*t*), *h*_*ar*_(*t*)) can be written as

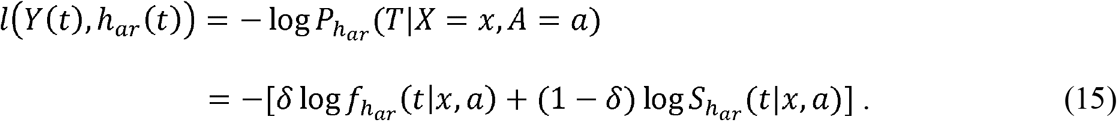

In general, using only the samples at risk with treatment status *A*, we define the loss function

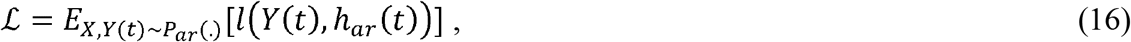

A treatment-specific hazard function 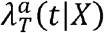 can be estimated by minimizing the following empirical risk function:

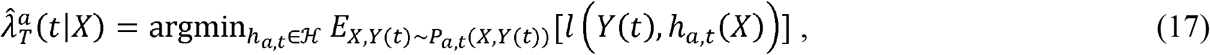

where ℋ is the hypothesis class or hazard function domain, and

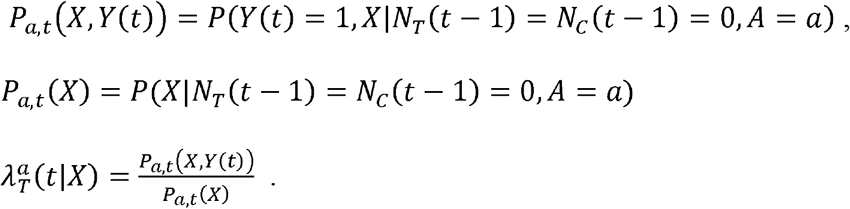

If the loss function is the log-likelihood loss function, then minimization (17) corresponds to optimizing the likelihood of the hazard.

*P*_*a,t*_ (*X*) is often called observational (at risk) covariate distribution. The distribution *P*_*a,t*_ (*X*) varies over time. Hence, treatment assignment will bias the estimation of the treatment effect. The ideal case for estimating the treatment effect is optimizing the loss function over the population at baseline. However, covariate shift in observational studies will exist and complicate the circumstance. Denote the marginal distribution of the covariates *X* at the baseline as *X*_0_ ∼ *P*_0_(*X*), three major sources will cause the changes of observational covariates distribution from baseline distribution *P*_0_ (*X*) to *P*_*a,t*_(*X*), including the treatment assignment bias and confounding, censoring and event-induced shifts [10].

The first source comes from treatment assignment bias and confounding. As discussed above, the observation covariate (at risk) distribution *P*_*a,t*_(*X*) depends on the assignment of the treatment. If in practice, the treatment violates the complete random assumption, then the covariate distribution will not be equal to the baseline covariate distribution *P*_0_ (*X*).

The second source comes from censoring. The treatment specific censoring hazard may depend on the covariates and treatment, i.e., λ_*C*_ (*t* | *a,x*) is a function of treatment *a* and covariates *X*. Censoring bias may cause the changes of populations at risk. In other words,

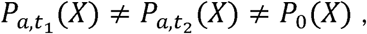

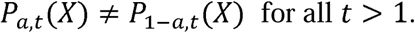

The third source comes from time-dependent effect and effect-induced covariate shifts. Both natural and vaccine immunity wanes over time. The effects of vaccine decrease when the time elapsed since vaccination. For example, the emergence of hypermutated new virus variants may cause escape from vaccine–induced immune responses. The gut microbiome in an individual’s host cells changes over time, which may cause shifts in the covariates in the population and even affect the immune responses to vaccination.

The importance weighting techniques were used to reweight the empirical risk. The survITE model [10] uses weight defined as the ratio of the target density *P*_0_ (*X*) and observes density distribution *P*_*a,t*_(*X*):

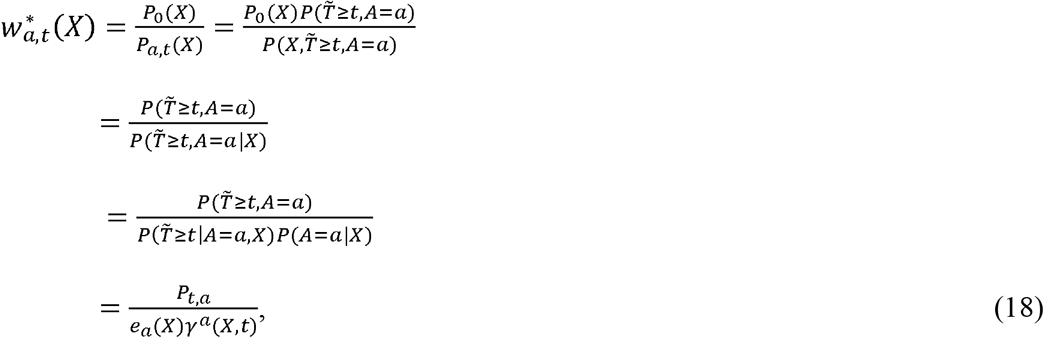

where

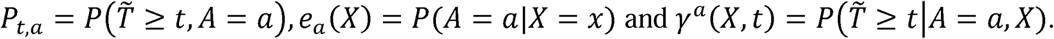

*e*_*a*_ (*X*) is often called the propensity score, and γ^*a*^ (*X,t*) is the probability to be at risk. Since the hazard parameter λ^*a*^ (*t* | *x*) and risk probability γ^*a*^ (*X,t*) are unknown, the weights are also unknown. For large,*t* survITE also requires weight 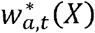 to satisfy the following normal distribution [10]:

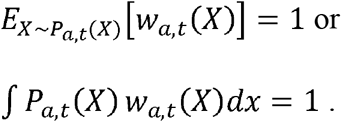

Therefore, we define the weight distribution as

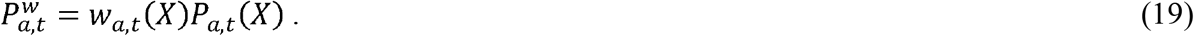

Domain-invariant representation is a useful technique to adapt the domain. Define a representation function learned by neural networks [14]: Ф (*x*). 𝒳 ⟶ *R* The time-varying hazard estimator 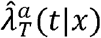 is a function of representation: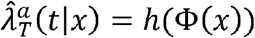. We require therepresentation Φ (*x*) should minimize the integral probability metric (IPM) between covariate distributions at baseline *P*_0_ and 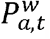. According to Fredrik et al. [15-16], there is an upper bound for the counterfactual generalization error of a representation function, Φ hence the optimization of the model would be minimization of the upper bound or loss function.

Next, we will introduce loss function and use it for estimation 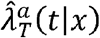

### Loss Function

Summarizing the analysis above, we introduce loss function for estimating individualized treatment effects from observational studies. The loss function borrowed from Cyrth et al. [10] is:

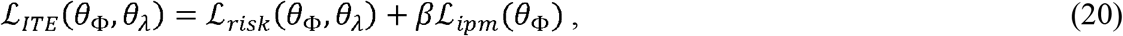

where

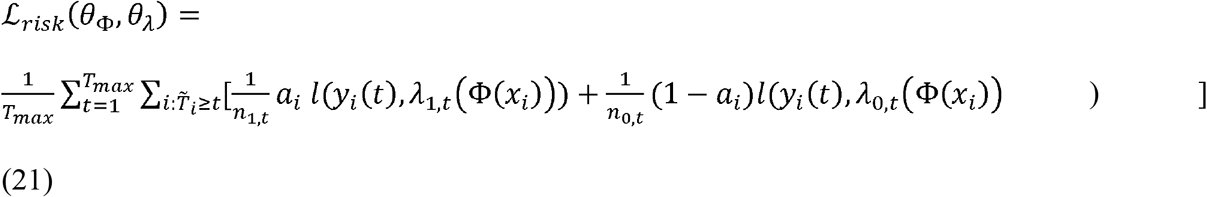

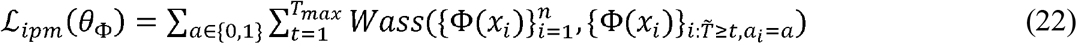

ϕ (*x*_*i*_)=*NN* (*x*_*i*_, θ Φ),*NN* (.,.)is a neural network function implementing Φ (*x*_*i*_), λ_*a,t*_ (Φ (*x*_*i*_))= *NN*(Φ (*x*_*i*_), *a, t, θ*_*λ*_) *NN* represented neural networks implementing λ_*a,t*_ (ϕ (*x*_*i*_)),

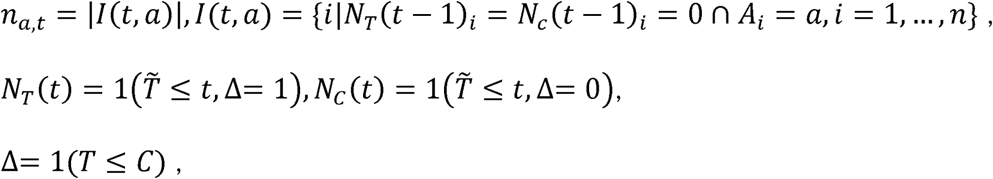

*Wass* (.)the Wasserstein distance which measures the difference of the distributions of *P*_*O*_ and 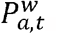.

The parameters *θ*_*Φ*_, *θ*_*λ*_ are the trained and estimated for the representation function Φ(x) and the hazard function. *λ*_*a,t*_ (x) can be estimated by minimizing the loss function ITE *ℒ*_*ITE*_(*θ*_*Φ*_, *θ*_*λ*_)

## Data Source

We apply the proposed model to estimate COVID-19 vaccination effectiveness in reducing break-through infection risk. Analysis cohort is obtained from Optum Electronic Health Records (EHR) research database. The Optum Research Database is an administrative claims database containing data from 160 million individuals and electronic health records for more than 80 million individuals nationally [17].

Laboratory and immunization data in Optum research database are manipulated. The COVID-19 infection is defined as SARS-CoV-2 laboratory test positive result. Break-through infection is defined as COVID-19 infection after patients get their second dose of vaccine. To analysis specific COVID-19 variant, the specimen collection dates restricted between June 1st 2021 and November 30th 2021, the period with Delta variant mainly spread in the United States. Four vaccine types mRNA-BNT162b2 (Pfizer–BioNTech), mRNA-1273 (Moderna), AZD-1222 (AstraZeneca) and Ad26.COV2. S (Johnson & Johnson–Janssen) - are recorded in the data, and the overall vaccine efficacy is estimated for all kinds of vaccine.

The demographic variables controlled in the model include age, gender, race, ethnicity and geographic region.

## Results

### Study Population

Data of patients who have COVID-19 test in the period from June 1st 2021 to November 30th 2021 were extracted from Optum research database. After data clean and manipulation, there were 957, 613 patients remaining in the dataset. In the 957, 613 patients, 127, 313 patients were fully vaccinated and 830, 300 patients did not have any dose of COVID-19 vaccines. In order to obtain balanced dataset for each treatment arms (fully vaccine vs non-vaccine), we randomly sampled 127, 313 patients in non-vaccine group. The final analyzed cohort contained 254, 626 patients. The demographic characteristics of the cohort is shown in Table 1.

**Table 1.** Demographic characteristics of the cohort.

| Age Group | Number of Individuals | Gender | Number of Individuals |
| --- | --- | --- | --- |
| < 12 yr | 27,325 (10.73%) | Female | 143,163 (56.22%) |
| 12-17 yr | 20,995 (8.25%) | Male | 111,463 (43.78%) |
| 18-34 yr | 51,503 (20.23%) | Ethnicity | Number of Individuals |
| 35-49 yr | 44,936 (17.65%) | Hispanic | 20,189 (7.93%) |
| 50-64 yr | 53,966 (21.19%) | Non-Hispanic | 186,639 (73.3%) |
| >=65 yr | 55,901 (21.95%) | Unknown | 47,798 (18.77%) |
| Race | Number of Individuals | COVID Infection Status | Number of Individuals |
| Africa American | 20,999 (8.25%) | Positive | 29,211 (11.47%) |
| Asian | 6,009 (2.36%) | Negative | 225,415 (88.53%) |
| Caucasian | 141,764 (55.68%) | Fully vaccine | 127,313 (50%) |
| Other/Unknown | 85,854 (33.72%) | Non-Vaccine | 127,313 (50%) |
| Geographic Region |  | South | 29,992 (11.78%) |
| Midwest | 80,687 (31.69%) | West | 25,563 (10.04%) |
| Northeast | 52,289 (20.54%) | Other/Unknown | 66,095 (25.96%) |

### Vaccine Effectiveness against Break-through Infection

Daily vaccine effectiveness was estimated by three models, survITE model [10], COX regression model [11] and RSF model [12]. The assumed follow-up time is 180 days (from June 1st 2021 to November 30th 2021).

This research also compared the survival probabilities and cumulative hazard functions between vaccine and non-vaccine groups. Since the survival probability and hazard function at each time point cannot be obtained by the package from COX regression model [11], the results of survITE model [10] and RSF model [12] were shown.

Figure 1 demonstrated the estimated vaccine effectiveness against break-through infection at different time points from three models, survITE model [10], COX regression model [11] and RSF model [12]. The mean vaccine effectiveness obtained by survITE model is 0.49, with maximum value at time point = 73 days, then vaccine effectiveness fell down with time passed. The mean vaccine effectiveness obtained by COX regression model is 0.88, with maximum value at time point = 29 days, then vaccine effectiveness slowly fell down with time passed. The mean vaccine effectiveness obtained by RSF model is 0.61, with maximum value at time point = 101 days, then vaccine effectiveness fell down with time passed. Figure 1 showed that the vaccine effectiveness estimated by the Cox regression model was much higher than the estimated by two other methods.

**Figure 1.**
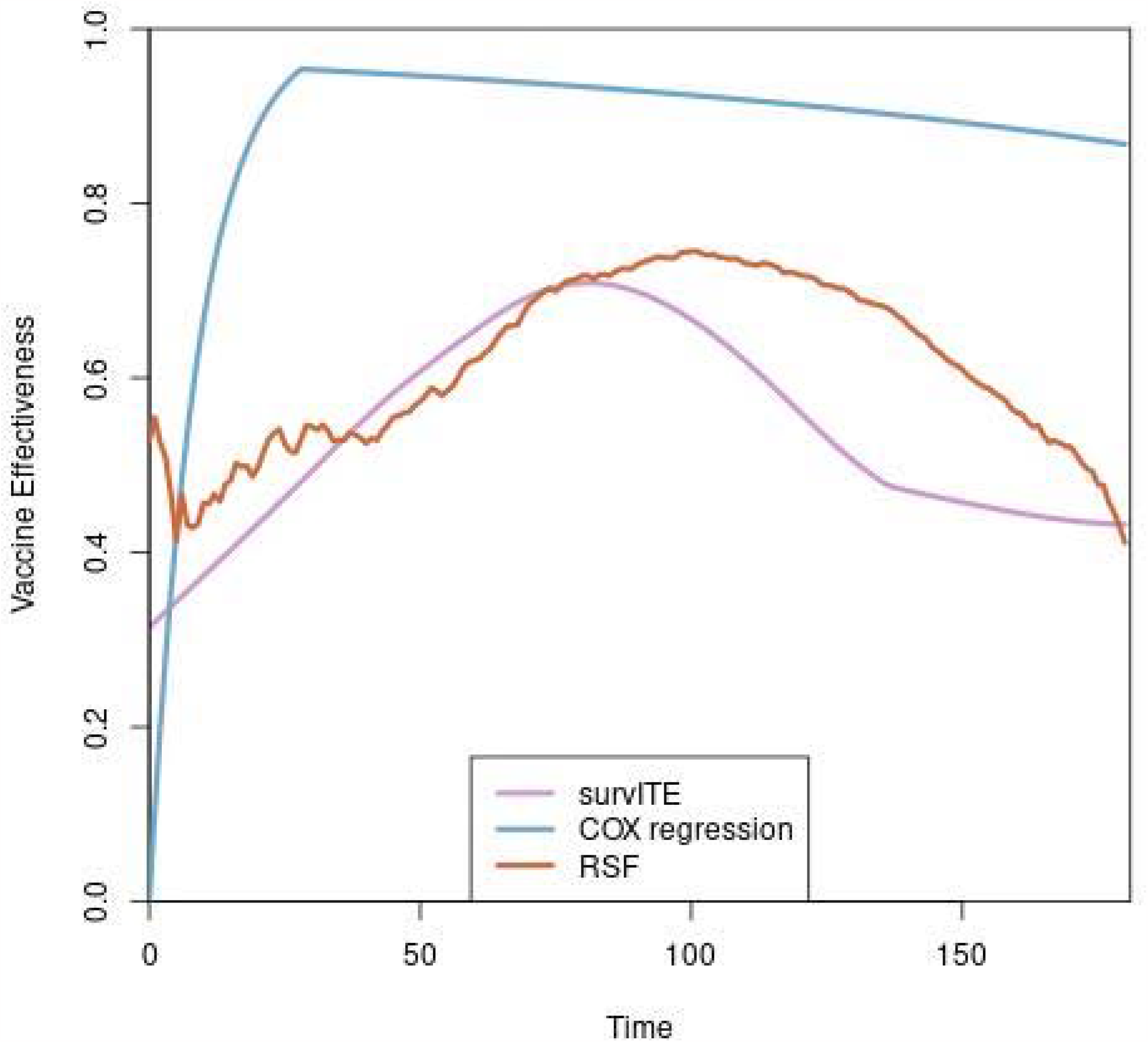
Vaccine effectiveness comparison among survITE model, Cox regression model and RSF model.

Figure 2 shows the survival probabilities of break-through infection obtained by proposed model at different time points, here vaccine and non-vaccine groups were plot separately. Figure 2 shows that survival probability of break-through infection for vaccine group was higher than non-vaccine group.

**Figure 2.**
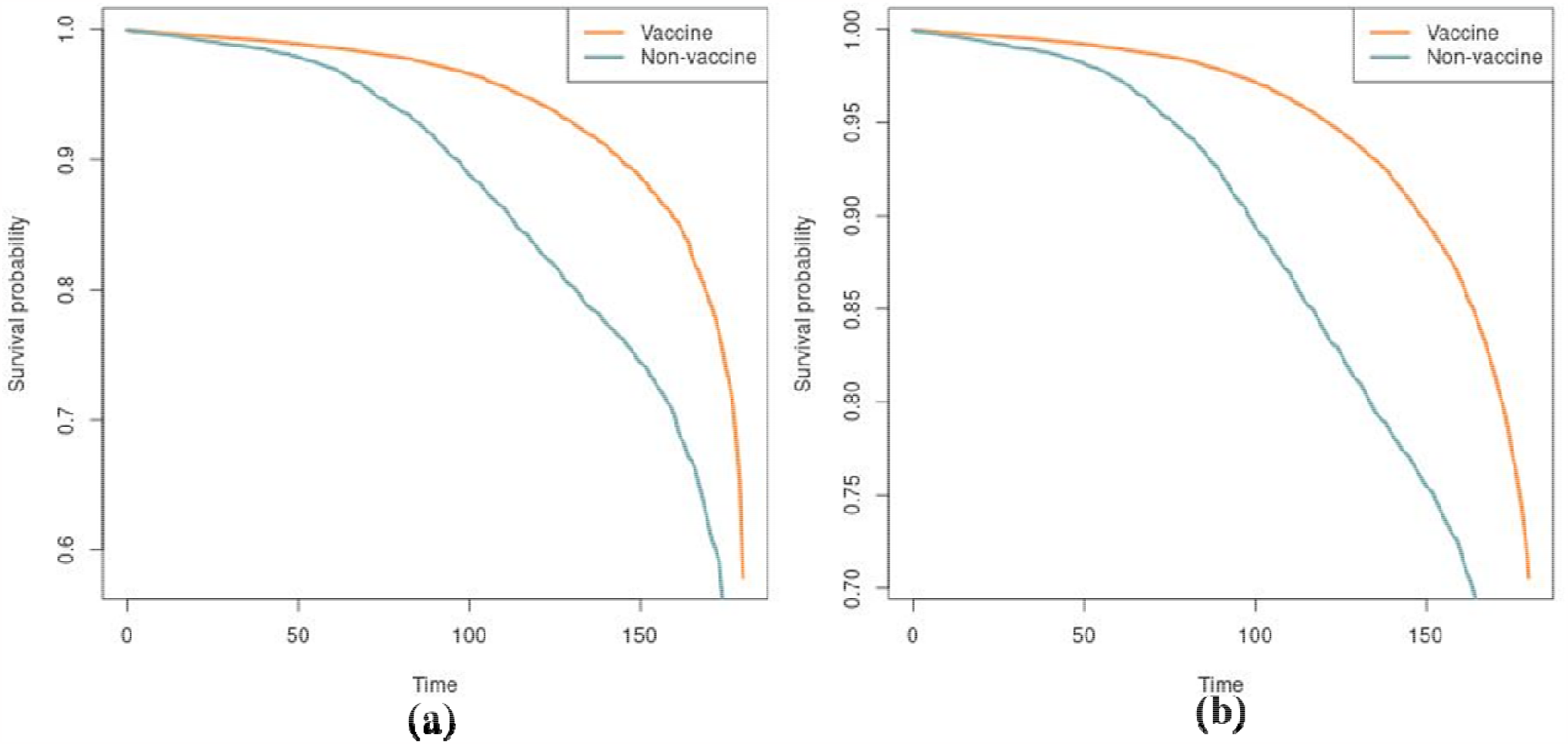
Survival probabilities at different time points estimated by survITE model (a) and RSF model (b) for vaccine and non-vaccine groups.

Figure 3 plotted the cumulative hazard function curves at different time points estimated by the survITE model (a) and RSF model (b) for the vaccine and non-vaccine groups. Figure 3 showed that difference in the cumulative functions between the vaccine and non-vaccine groups estimated by the SurvITE model was smaller that that estimated by the RSF models. Since the software for Cox regression model does not provide calculation of the survival probability and hazard functions, Figure 3 did not include the results of the Cox regression models.

**Figure 3.**
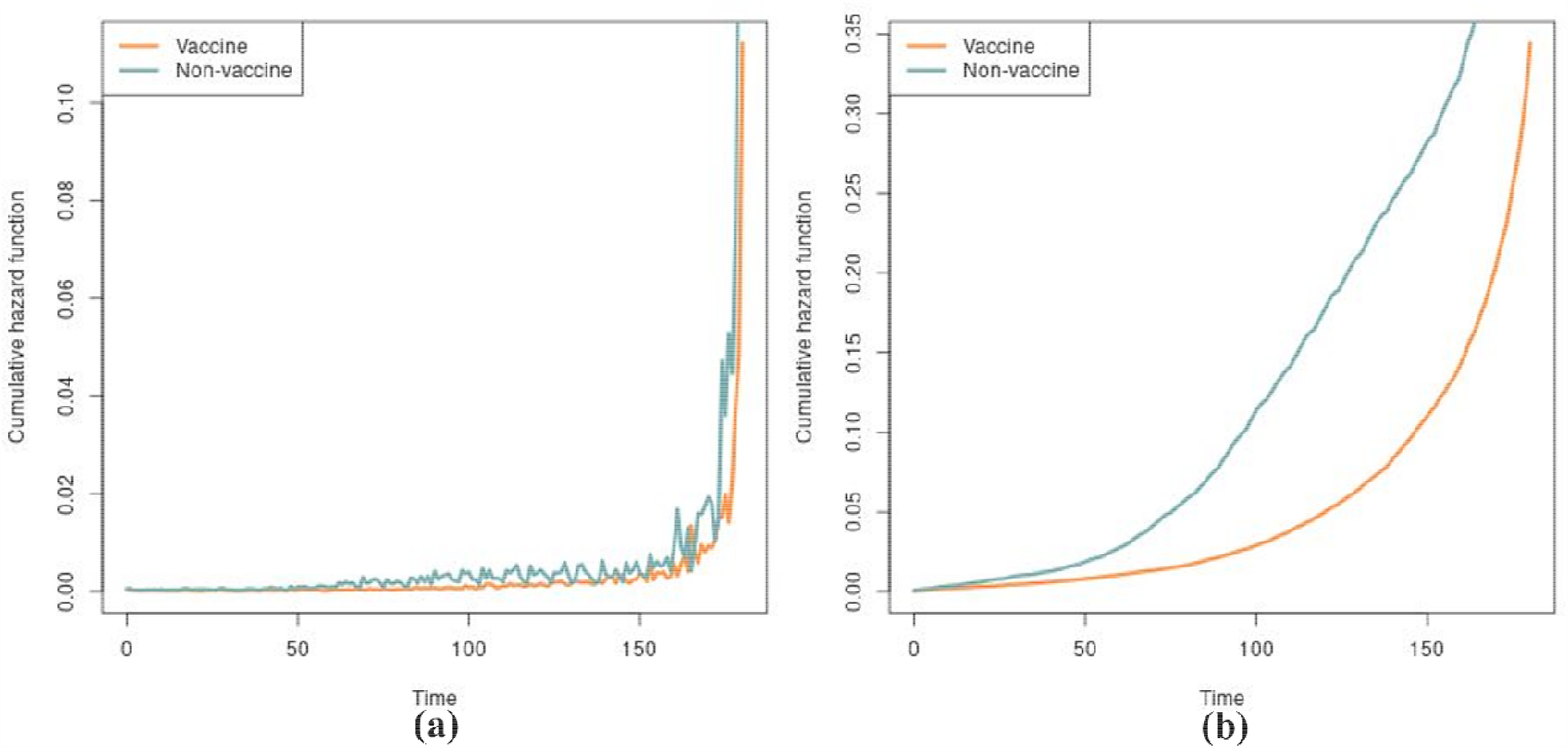
Cumulative hazard functions at different time points estimated by survITE model (a) and RSF model (b) for vaccine and non-vaccine groups.

Figures 4 and 5 illustrated the survival probabilities of break-through infection for vaccine and non-vaccine groups estimated by the survITE and RSF models, respectively. The difference in estimations of survival probabilities between the survITE and RSF models was small.

**Figure 4.**
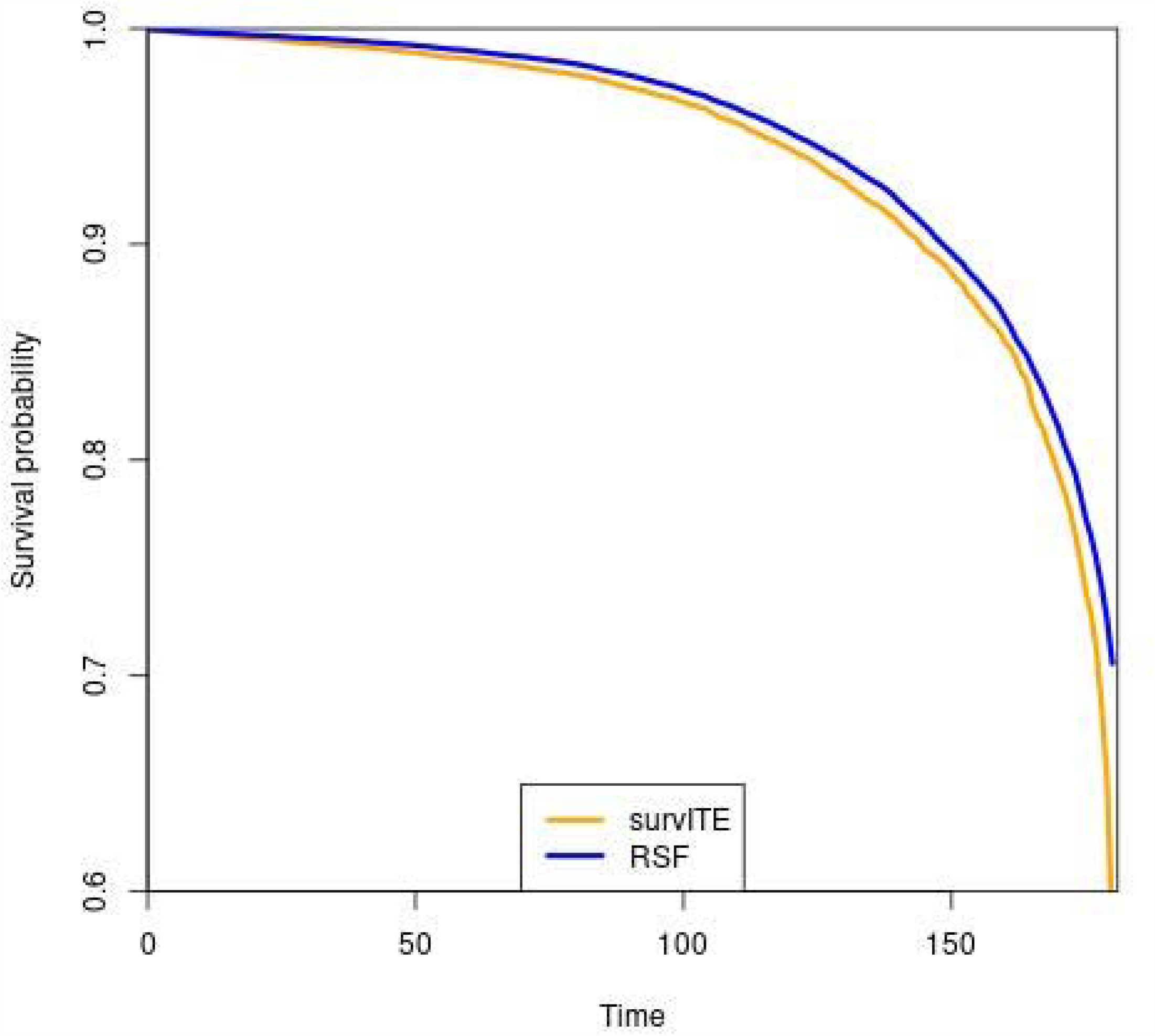
The survival probabilities of vaccine group at the different time points estimated by two models: survITE and RSF models.

**Figure 5.**
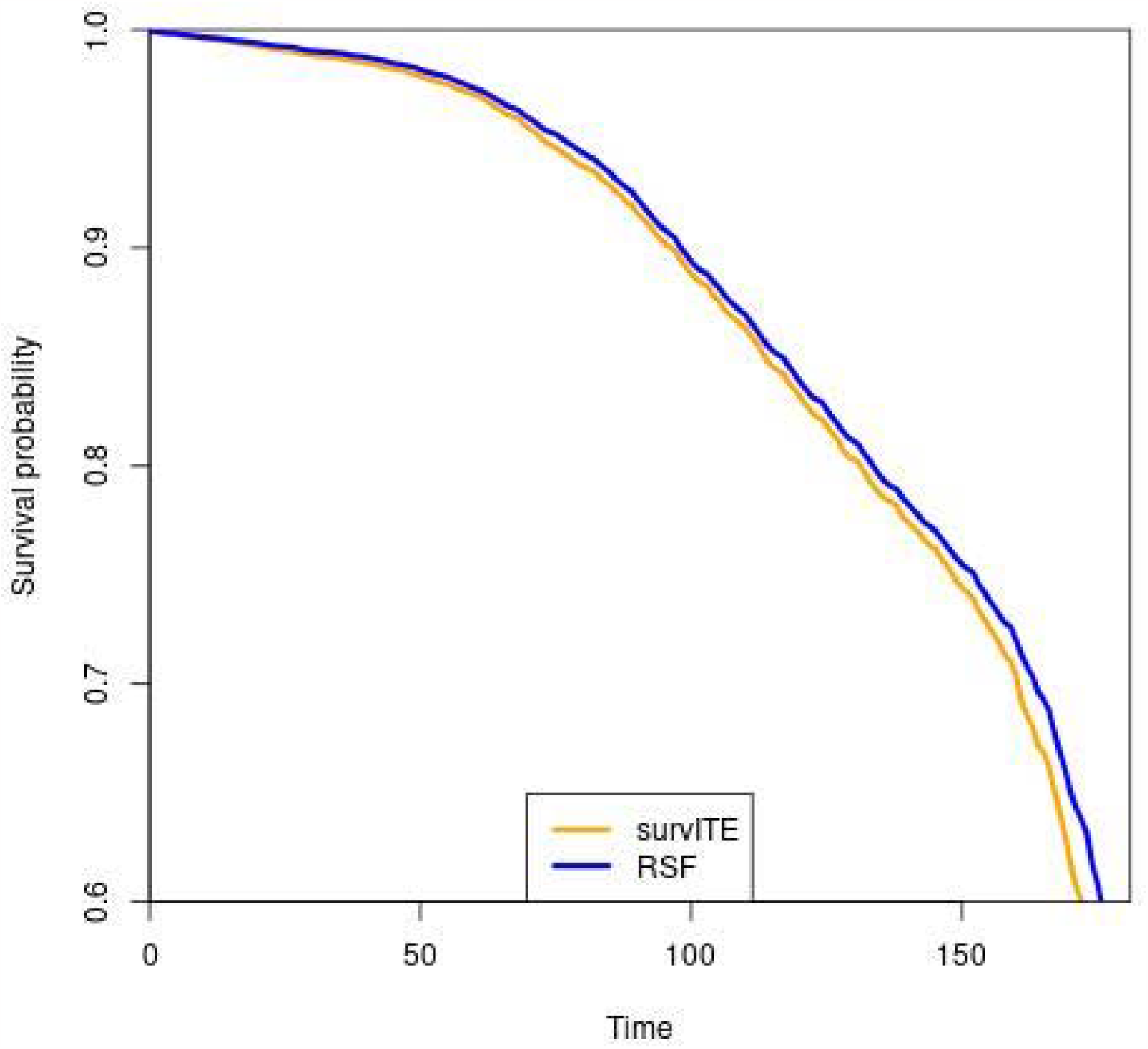
The survival probabilities of non-vaccine group at the different time points estimated by two models: survITE and RSF models.

Figures 6 and 7 illustrated the cumulative hazard functions of break-through infection for vaccine and non-vaccine groups, estimated by the survITE and RSF models, respectively. We observed from Figures 6 and 7 that the difference in the estimated hazard function between the survITE and RSF models in the vaccine group was smaller than that in the non-vaccine group.

**Figure 6.**
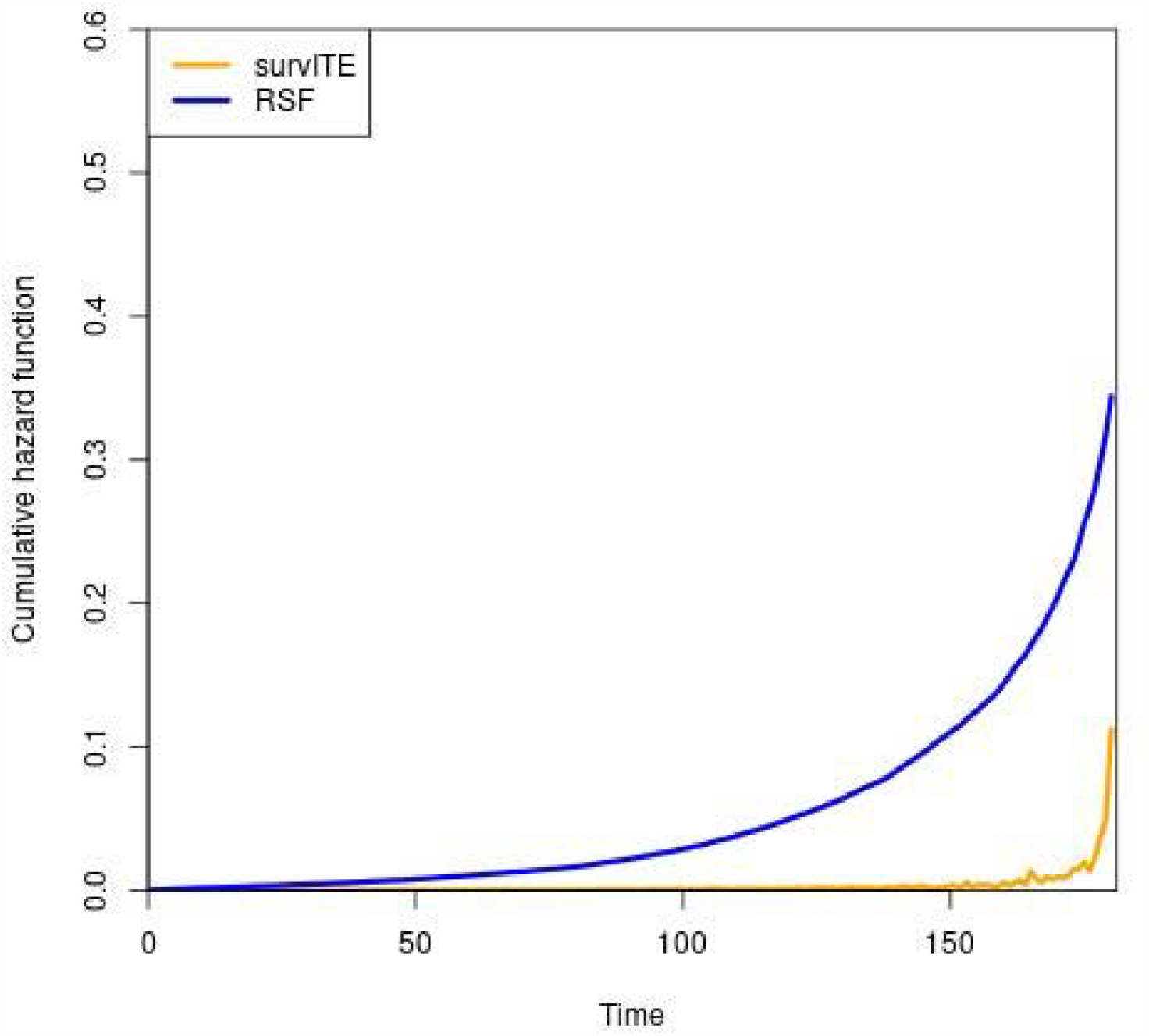
The cumulative hazard functions of the vaccine group at the different time points estimated by two models: survITE model and RSF model.

**Figure 7.**
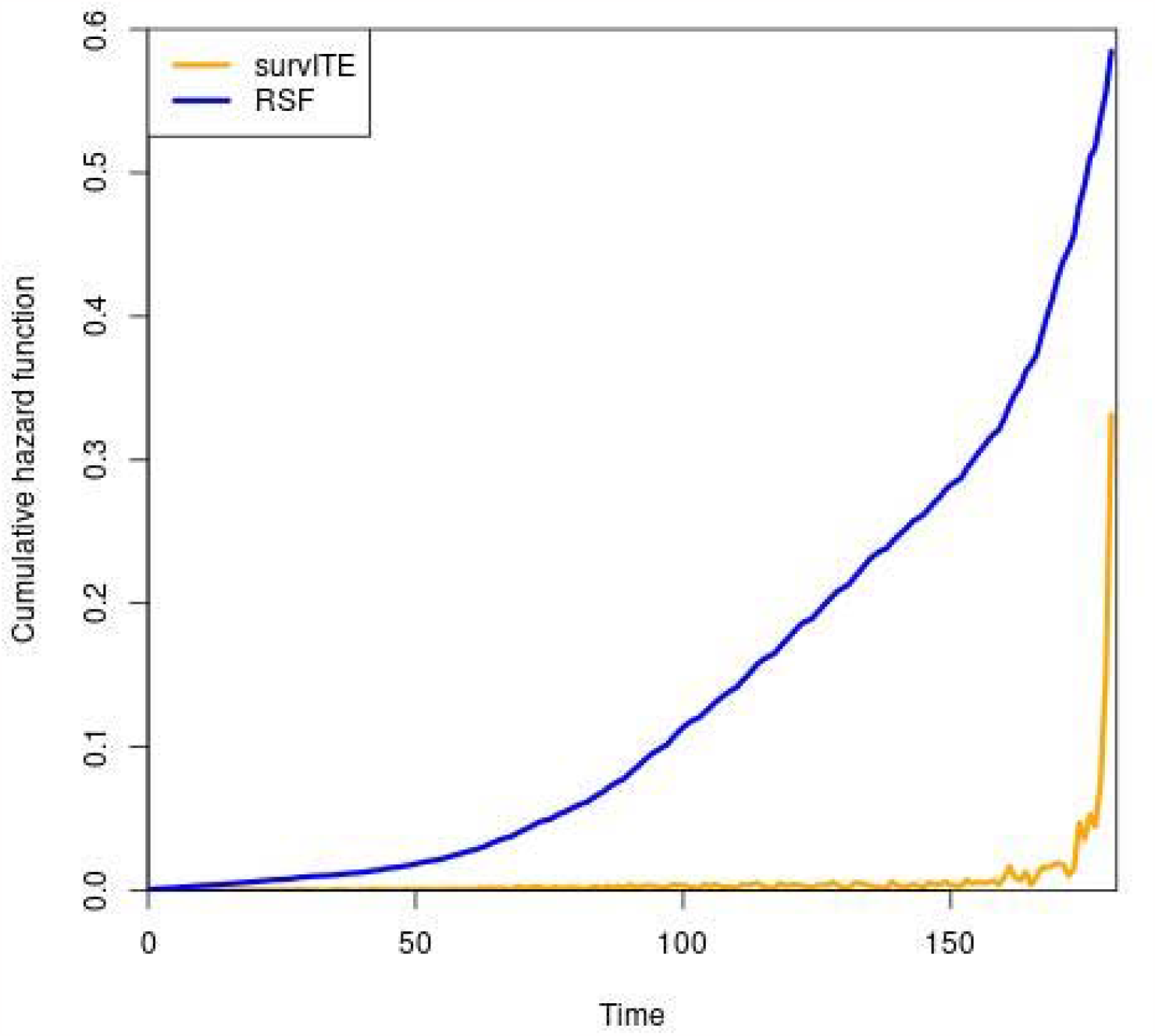
The cumulative hazard functions of the non-vaccine group at the different time points estimated by two models: survITE model and RSF model.

## Discussion

In this report, the EHR dataset was used for the estimation of COVID-19 vaccine effects. The Optum EHR dataset was cleaned and manipulated. Since there were several different types of coronaviruses, to reduce the bias caused by virus type, the observation time interval was restricted into the period from June 1^st^, 2021 to November 30^th^, 2021. And since the vaccine types in dataset were not indicated, the VE estimation included mRNA-BNT162b2 (Pfizer– BioNTech), mRNA-1273 (Moderna), AZD-1222 (AstraZeneca) and Ad26.COV2. S (Johnson & Johnson–Janssen).

The estimated daily VE values from the three models, survITE model, Cox regression model and RSF model had revealed the changing trend of VE with time. The three curves showed similar trend in the beginning of observational time interval, the VE increased after the injection of vaccine, but the results of three models showed different speed for VE increasing. Cox regression model showed that the VE increased rapidly and reached a peak in a very short time period (29 days) after fully vaccinated, and the increasing speed for the other two models (survITE: 73 days, RSF: 101 days) is relatively low. After reached a peak, VE started to decrease, and both survITE model and RSF model more rapidly decrease than Cox regression model. The changing trends of VE for three models approximately followed other vaccine effectiveness studies [6-7].

The selection bias and covariate shift exist in observational data research, and the dataset used in this research also encountered with these problems. Since survITE model applied counterfactual reasoning by representation learning from demographic variables of patients, which decreased the covariate shift and selection bias in the real-world problems, the calculation of hazard function would be closer to the real value. The Cox regression model was based on Cox PH model. Althouth it introduced the dynamic elements in the calculation, it still could not eliminate the selected bias and variable shift generated from observational data and overestimated the VE. RSF model was a forest ensemble learner for analysis of right-censored survival data [18]. And it seemed that the estimated VE from RSF model was lower than Cox regression model, but higher than survITE model. This may imply that the RSF model could eliminate some effects due to selection bias and variate shift, but still was not be able to unbiasedly estimate the VE.

Afterall, from the results of three models, we observed that based on Optum EHR dataset, COVID-19 vaccine increased the survival probability of an individual from getting breakthrough infection and the VE of COVID-19 against breakthrough infection increased after injection of vaccine. However, selection bias and variable shift widely exist. Most statistical methods for VE estimation are based association. Vaccine effectiveness (VE) estimation is a heterogeneous treatment effects from time-to event data. It consists of two parts: (1) estimating treatment effects for potential binary or continuous outcomes and (2) predicting survival outcomes. Three factors: (1) confounding, (2) censoring and (3) a variety of covariate shift will bias the estimation. The VE estimation involves hazard and survival functions which are determined by the dynamics of the underlying stochastic processes. The complex stochastic processes in turn lead to the shift of variates. These features make the time to event outcomes significantly different from the regression targets which leads to modeling the time-to-event outcomes difficult and the biased VE estimation.

Widely used Cox regression model is not causally interpretable even if in a randomized survival study [19], [20]. The HR obtained from the Cox regression model cannot be given a causal interpretation. The shift of covariates will change the selection and change the hazard ratios, which in turn change the VE. The VE is not only measured by the vaccine, but also by the stochastic virus, biologic properties of the individuals, other public health interventions, and environments. Therefore, the Cox regression model overestimated the VE.

In contrast, the SurvITE formalized the survival treatment effect problem in terms of empirical risk minimization, used neural network-based model for balanced representations and designed specific loss functions with some regularizations for estimation of treatment-specific target parameters (hazard and survival functions) which took mitigate the impact of shifts of covariates on the parameter estimation into account. Thereforfe, the VE estimated by the SurvITE was less biased than the COX regression and RSF models.

Limitations of SurvITE are as follows. The SurvITE still relies on a set of strong assumptions which have not been validated in both theory and practice. Random censoring is an additional assumption added to the standard ‘no hidden confounders’ assumption in classical treatment effect estimation, which may invalid any meaningful causal estimation of heterogeneous VE estimation.

## Data availability

All datasets used in this work are available on request.

## Code availability

The code for estimation of vaccine effects will be submitted to github very soon.

## Reference

[1] CDC (Centers for Disease Control and Prevention) COIVD Data Tracker. Internet Archive. https://www.cdc.gov/coronavirus/2019-ncov/covid-data/covidview/index.html. xPublished 2023. Accessed July 31, 2023.

[2] Lin D, Zeng D, Gilbert PB. Evaluating the long-term efficacy of coronavirus disease 2019 (COVID-19) vaccines. Clin Infect Dis. 2021;73(10):1927–1939. doi: 10.1093/cid/ciab226.

[3] Biswas N, Mustapha T, Khubchandani J, Price JH. The nature and extent of COVID-19 vaccination hesitancy in healthcare workers. J Community Health. 2021;46(6):1244–1251. doi: 10.1007/s10900-021-00984-3.

[4] Lipsitch M, Dean NE. Understanding COVID-19 vaccine efficacy. Science. 2020;370(6518): 763–765. doi: 10.1126/science.abe5938.

[5] Knoll MD, Wonodi C. Oxford–AstraZeneca COVID-19 vaccine efficacy. Lancet. 2021;397(10269): 72–74. doi: 10.1016/S0140-6736(20)32623-4.

[6] Shinde V, Bhikha S, Hoosain Z, et al. Efficacy of NVX-CoV2373 covid-19 vaccine against the B.1.351 variant. N Engl J Med. 2021;384(20):1899–1909. doi: 10.1056/NEJMoa2103055.

[7] Heath PT, Galiza EP, Baxter DN, et al. Safety and efficacy of NVX-CoV2373 covid-19 vaccine. N Engl J Med. 2021;385(13):1172–1183. doi: 10.1056/NEJMoa2107659.

[8] Nikas A, Ahmed H, Zarnitsyna VI. Estimating waning of vaccine effectiveness: A simulation study. Clin Infect Dis. 2023;76(3):479–486. doi: 10.1093/cid/ciac725.

[9] Durham LK, Longini IM, Halloran ME, Clemens JD, Azhar N, Rao M. Estimation of vaccine efficacy in the presence of waning: Application to cholera vaccines. Am J Epidemiol. 1998;147(10):948–959. doi: 10.1093/oxfordjournals.aje.a009385.

[10] Curth A, Lee C, van der Schaar M. SurvITE: Learning heterogeneous treatment effects from time-to-event data. arXiv.org. 2022.

[11] Lin D, Gu Y, Wheeler B, et al. Effectiveness of Covid-19 vaccines over a 9-month period in North Carolina. New England Journal of Medicine, 2022;386(10): 933–941. doi: 10.1056/NEJMoa2117128

[12] Ishwaran H, Kogalur UB, Blackstone EH, Lauer MS. Random survival forests. The annals of applied statistics. 2008;2(3):841–860. doi: 10.1214/08-AOAS169.

[13] Kvamme H, Borgan Ø. Continuous and discrete-time survival prediction with neural networks. Lifetime Data Anal. 2021;27(4):710–736. doi: 10.1007/s10985-021-09532-6.

[14] Johansson FD, Sontag D, Ranganath R. Support and invertibility in domain-invariant representations. arXiv.org. 2019.

[15] Johansson FD, Shalit U, Sontag D. Learning representations for counterfactual inference. arXiv.org. 2018.

[16] Shalit U, Johansson FD, Sontag D. Estimating individual treatment effect: Generalization bounds and algorithms. arXiv.org. 2017.

[17] Toth PP, Hull M, Granowitz C, et al. Real-world analyses of patients with elevated atherosclerotic cardiovascular disease risk from the Optum Research Database. Future Cardiology, 2020;17(4):743–755. doi: 10.2217/fca-2020-0123.

[18] Ishwaran, Hemant, and Udaya B. Kogalur. “Consistency of random survival forests.” Statistics & probability letters, 2010;80(13-14): 1056–1064. doi: 10.1016/j.spl.2010.02.020.

[19] Aalen OO, Cook RJ, Røysland K. Does Cox analysis of a randomized survival study yield a causal treatment effect? Lifetime Data Analysis. 2015, 21(4): 579–593.

[20] Martinussen T. Causality and the Cox regression model. Annu. Rev. Stat. Appl. 2022. 9:249–59.

