## Supplementary material for "Causal Inference for Estimation of Vaccine Effects from Time-to-Event Data": Figure S1

**Supplemental Files**

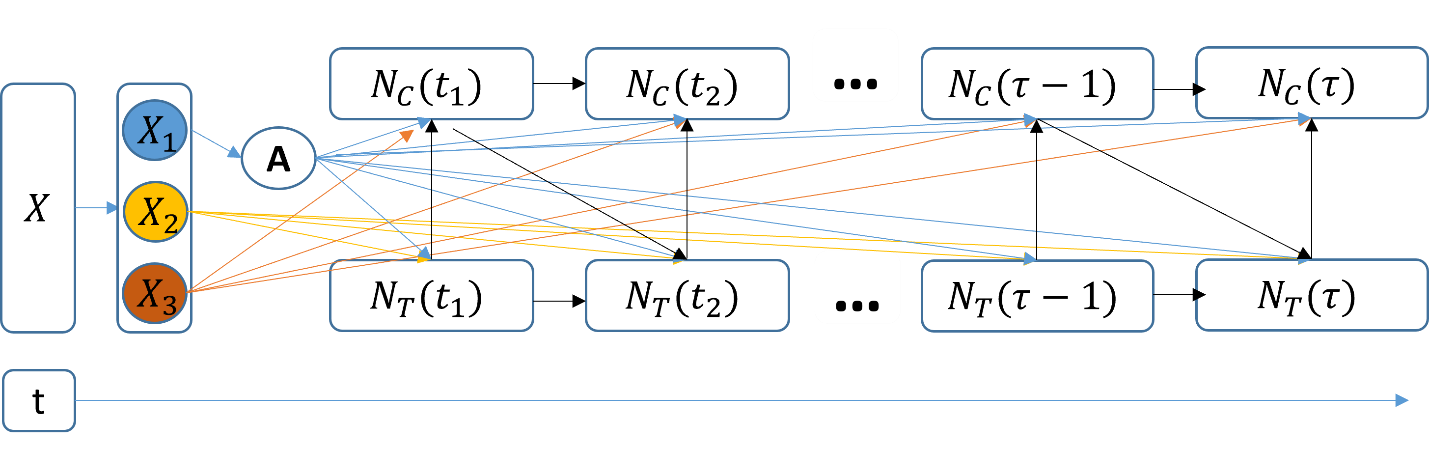

**Figure S1**. Generally directed acyclic graphs (DAGs) for data generation of time-to-event data. $X_{1}, X_{2}, X_{3}$ are subsets of covariates, which stands for covariates shift sources affect treatment

selection, event times and information censoring.
